# Establishing and characterising large COVID-19 cohorts after mapping the Information System for Research in Primary Care in Catalonia to the OMOP Common Data Model

**DOI:** 10.1101/2021.11.23.21266734

**Authors:** Edward Burn, Sergio Fernández-Bertolín, Erica A Voss, Clair Blacketer, Maria Aragón, Martina Recalde, Elena Roel, Andrea Pistillo, Berta Raventós, Carlen Reyes, Sebastiaan van Sandijk, Lars Halvorsen, Peter R Rijnbeek, Talita Duarte-Salles

## Abstract

**Background:** Few datasets have been established that capture the full breadth of COVID-19 patient interactions with a health system. Our first objective was to create a COVID-19 dataset that linked primary care data to COVID-19 testing, hospitalisation, and mortality data at a patient level. Our second objective was to provide a descriptive analysis of COVID-19 outcomes among the general population and describe the characteristics of the affected individuals.

**Methods:** We mapped patient-level data from Catalonia, Spain, to the Observational Medical Outcomes Partnership (OMOP) Common Data Model (CDM). More than 3,000 data quality checks were performed to assess the readiness of the database for research. Subsequently, to summarise the COVID-19 population captured, we established a general population cohort as of the 1^st^ March 2020 and identified outpatient COVID-19 diagnoses or positive test results for SARS-CoV-2, hospitalisations with COVID-19, and COVID-19 deaths during follow-up, which went up until 30^th^ June 2021.

**Findings:** Mapping data to the OMOP CDM was performed and high data quality was observed. The mapped database was used to identify a total of 5,870,274 individuals, who were included in the general population cohort as of 1^st^ March 2020. Over follow up, 604,472 had either an outpatient COVID-19 diagnosis or positive test result, 58,991 had a hospitalisation with COVID-19, 5,642 had an ICU admission with COVID-19, and 11,233 had a COVID-19 death. People who were hospitalised or died were more commonly older, male, and with more comorbidities. Those admitted to ICU with COVID-19 were generally younger and more often male than those hospitalised in general and those who died.

**Interpretation:** We have established a comprehensive dataset that captures COVID-19 diagnoses, test results, hospitalisations, and deaths in Catalonia, Spain. Extensive data checks have shown the data to be fit for use. From this dataset, a general population cohort of 5.9 million individuals was identified and their COVID-19 outcomes over time were described.

**Funding:** Generalitat de Catalunya and European Health Data and Evidence Network (EHDEN).

## Introduction

Spain has been one of the European countries hit hardest by the ongoing Coronavirus disease 2019 (COVID-19) pandemic. The first COVID-19 cases in Spain were identified in late February 2020, and by the 1^st^ May of that year there had been more than 32,000 COVID-19 deaths in the country.^1^ Further waves of infections have since followed and although the advent of effective vaccines has dramatically improved the outlook, COVID-19 cases continue to accrue and the effects of the disease for many of the people previously infected are likely to be long-lasting.

Similar to many European countries, Spain has a universal coverage healthcare system with general practitioners (GPs) acting as the gatekeepers to care.^2^ This role has largely been maintained during the COVID-19 pandemic. In particular, clinical diagnoses by primary care professionals played an important role during the first wave of COVID-19 in Spain, with the use of SARS-CoV-2 testing initially restricted to the most severe cases, such as those hospitalised and groups considered to be at particularly high-risk, such as care home residents.^3^

In such a context, primary care records can provide an important foundation for COVID-19 research, particularly when linkage to testing, hospitalisation, and mortality data is available. The Information System for Research in Primary Care (SIDIAP; www.sidiap.org) is a primary care records database covering approximately 80% of the population of Catalonia. The provenance of the data has been well documented, and the population captured have been found to be representative in terms of geography, age, and sex.^4^ Data from SIDIAP has previously been used for in a wide range of epidemiological research studies, including COVID-19 related research.^5–8^ Individual-level linkage of hospital and regional mortality data has previously been established for SIDIAP, and now further linkage to SARS-CoV-2 test results is also possible.

Our first aim was to establish a SIDIAP COVID-19 dataset that could be used to generate reliable evidence relating to the pandemic. To facilitate international collaboration, we set out to conform to the Observational Medical Outcomes Partnership (OMOP) Common Data Model (CDM).^9^ Our second aim was to summarise the occurrence of COVID-19 outcomes observed and describe the characteristics of those affected.

## Methods

### Overview

Primary care data collected in SIDIAP between 1^st^ January 2006 (when data became available) and 30^th^ June 2021 (the last available date of data collection) was linked, at a patient-level, to COVID-19 testing, hospitalisation, and mortality data. This data was mapped to the OMOP CDM following an extract, transform, and load process which we first describe below. Using this mapped data, a cohort of the general population was followed up from 1^st^ March 2020, with COVID-19 outcomes (outpatient COVID-19 diagnoses and positive tests, hospitalisations with COVID-19, and COVID-19 deaths) observed over follow-up until the 30^th^ June 2021.

### Mapping to the OMOP CDM: Extract, Transform, and Load

The extract, transform, and load (ETL) process was based on the approach put forward by the Observational Health Data Sciences and Informatics (OHDSI) community which involves four distinct steps: 1) designing the ETL, 2) creating the code mappings, 3) implementing the ETL, and 4) quality control to assess whether the database was fit for use. Any issues identified during quality control are addressed by updating the ETL where possible.^10^

#### Designing the ETL

The OHDSI White Rabbit tool was used to scan and characterise the source data.^11^ Based on this, a design was created using the Rabbit-in-a-Hat tool in which source data tables were mapped to the OMOP CDM person, observation period, visit occurrence, condition occurrence, procedure occurrence, drug exposure, measurement, observation, and death tables (see Supplementary Figure 1). The derived drug and condition era tables were also created.

#### Creating the code mappings

Mapping to the OMOP CDM requires mapping terminology to standard vocabularies in the OMOP Vocabularies.^11^ Example of code mappings are given in Table 1. SNOMED, for example, is a standard vocabulary for conditions, while RxNorm codes are a standard vocabulary for drug exposures.

**Table 1.** Example of mappings implemented in SIDIAP to the OMOP CDM

| Source Data |  |  | OMOP CDM |  |  |
| --- | --- | --- | --- | --- | --- |
| Table | Concept (Vocabulary) | Description | Table | Concept ID (Vocabulary) | Description |
| **COVID-19 Diagnoses** |  |  |  |  |  |
| *Diagnostics* (diagnoses) | "B34.2"<br>(ICD10 CM) | Coronavirus infection, unspecified site | Condition occurrence | 439676<br>(SNOMED) | Coronavirus infection |
| *Diagnostics* (diagnoses) | "B97.29"<br>(ICD10 CM) | Other coronavirus as the cause of diseases classified elsewhere | Condition occurrence | 4100065<br>(SNOMED) | Disease due to Coronaviridae |
| *Covid formulari seguiment* (COVID monitoring) | "INTERPRETACI O_COVID-19 confirmada greu"<br>(N/A) | COVID-19 confirmed (grave) | Condition occurrence | 37311061<br>(SNOMED) | Disease caused by 2019 novel coronavirus |
| **SARS-CoV-2 test results** |  |  |  |  |  |
| *PCR* | "Positiu"<br>(N/A) | Positive COVID19 PCR Result | Measurement | 37310255<br>(SNOMED) - 45884084<br>(LOINC) | Detection of 2019 novel coronavirus using polymerase chain reaction technique - Positive |
| **Symptoms** |  |  |  |  |  |
| *Diagnostics* (diagnoses) | "R06.02"<br>(ICD10 CM) | Shortness of breath | Condition occurrence | 312437<br>(SNOMED) | Dyspnea |
| *Covid formulari seguiment* (COVID monitoring) | "DISPNEA (*no greu*)" (N/A) | Dyspnea (not serious) | Condition occurrence | 312437 (SNOMED) | Dyspnea |
| **Comorbidities** |  |  |  |  |  |
| *Diagnostics* (diagnoses) | "E11.9" (ICD10CM) | Type 2 diabetes mellitus without complications | Condition occurrence | 4193704 (SNOMED) | Type 2 diabetes mellitus without complication |
| *Covid formulari seguiment* (COVID monitoring) | "DM" (N/A) | Diabetes Mellitus | Condition occurrence | 201820 (SNOMED) | Diabetes mellitus |
| **Medications** |  |  |  |  |  |
| *prescripcio* (prescription) | "656509" (AEMPS national code) | BRUFEN, 600 MG 40 COMPRIMIDOS, COMPRIMIDOS | Drug exposure | 19019073 (RxNorm) | Ibuprofen 600 MG Oral Tablet |

Mapping from source data to the Observational Medical Outcomes Partnership (OMOP) Common Data Model (CDM) requires source data to be mapped to standard concepts. For example, SNOMED provides standard concepts for the condition occurrence table. Here we provide some examples of the mappings used. AEMPS: *Agencia Española de Medicamentos y Productos Sanitarios*.

COVID-19 diagnoses and patient comorbidities could first be identified from the source table *diagnostics* (diagnoses) table, containing ICD-10 codes recorded during primary care interactions. An example of a COVID-19 diagnosis code mapping for this table was from the ICD-10 code B34.2 (“ Coronavirus infection, unspecified site”) to the OMOP concept id 439676 (“ Coronavirus infection”), while an example of a COVID-19 symptom code is the mapping of the ICD-10 code R06.02 (“ Shortness of breath”) to the concept id 312437 (“ Dyspnea”). Additional data specifically collected for people with COVID-19 was captured in the *covid formulari seguiment* (COVID monitoring) table. These data were not codified in the source table, but instead contained columns representing a concept specific to COVID-19 (with a value of 1 entered if it was present). As an example, a value of 1 in the column with the name *interpretacio COVID-19 confirmada greu* (COVID-19 – confirmed, serious) was mapped to the concept id 37311061 (“ Disease caused by severe acute respiratory syndrome coronavirus 2”). Likewise, a value of 1 the column “ anosmia” was mapped to the concept id 4185711 (“ Loss of sense of smell”).

Prescriptions of medications were identified from the source table *prescripcio* (prescription), with this information mapped to the drug exposure table of the OMOP CDM. To map each drug national code from AEMPS (*Agencia Española de Medicamentos y Productos Sanitarios*) in the source table to the best corresponding standard concept id in the OMOP CDM drug exposure table an intermediate table was used

SARS-CoV-2 test results were identified from a new source table, linked to SIDIAP patient data at the individual-level. These results were mapped to the measurement table in the OMOP CDM. Each polymerase chain reaction (PCR) test record in this source table were mapped to a measurement concept id of 586310 (“ Measurement of Severe acute respiratory syndrome coronavirus 2 (SARS-CoV-2) Genetic material using Molecular method”), while antigen tests were mapped to 37310257 (“ Measurement of Severe acute respiratory syndrome coronavirus 2 antigen”) and antibody tests to 37310258 (“ Measurement of Severe acute respiratory syndrome coronavirus 2 antibody”). If the result was coded as *Positiu* (positive) in the source table, the value as concept id was then set as the concept id of 45884084 (“ Positive”).

Lastly, hospitalisation data, from the conjunt mínim bàsic de dades de l’alta hospitalària (minimum basic set of hospital discharge data) collated by the Data Analysis Program for Health Research and Innovation (PADRIS) in Catalonia, was also linked at the individual-level. This dataset included both diagnosis and procedures registered during hospital admissions for all public and private hospitals in Catalonia. ICD-10 codes used to register diagnoses at hospitals were mapped to the OMOP CDM, as was done with the *diagnostics* table, with the procedure occurrence table in the CDM also populated.

#### Implementing the ETL

The local SIDIAP tables are in a MariaDB database, with the OMOP CDM v5.3.1 tables stored in a PostgreSQL database. Docker containers were used to host the full environment. SQL code was used for implementing the mapping in the MariaDB environment, with GitHub used for version control. After completing the mapping, CDM tables were migrated to their final PostgreSQL environment. A schematic of the extract, transform, and load (ETL) framework used is provided in Supplementary Figure 2.

#### Quality control

A range of database constraints defined by OHDSI were created in the Postgres database to prevent errors such as duplicate rows or unmatched id’s across interrelated tables. Data quality was also considered systematically using the Data Quality Dashboard (DQD) tool.^11,12^ This tool was run on the data after conversion to the OMOP CDM to test how well the resulting CDM instance complied with OHDSI standards. DQD runs a series of data quality checks that measure a database’s conformance to the model specifications, completeness of mapping to Standard Concepts, and plausibility of a select set of values. At a high-level, the tool works by applying data quality check types to applicable tables and fields in the CDM. The results of DQD were used to evaluate whether the database was fit for use. In total, 3,486 data quality checks were performed, any failed checks were reviewed, and the ETL was updated to address them where necessary.

### Summarising the occurrence of COVID-19 outcomes and describing the characteristics of those affected

#### Study population and follow-up

Individuals present in SIDIAP as of 1^st^ March 2020 were identified as the study population. Any individuals who had a clinical diagnosis or positive test result for SARS-CoV-2 between the 1^st^ January and 29^th^ February 2020 were excluded, as were any individuals in hospital on 1^st^ March 2020. These two exclusions were to ensure that the cohort identified from SIDIAP was representative of the general population at risk of subsequent incident COVID-19. Follow-up for this cohort began on 1^st^ March 2020 (the index date for all individuals) and ended on 30^th^ June 2021 (the last available date of data collection).

#### COVID-19 outcome cohorts

Four COVID-19 related outcomes were considered: outpatient COVID-19, hospitalised with COVID-19, ICU admission with COVID-19, and died with COVID-19. These outcome cohorts were not mutually exclusive.

An outpatient diagnosis of COVID-19 was identified on the basis of a compatible clinical code (with a broad definition used) or positive SARS-CoV-2 test (antigen or PCR), with no hospital admission with COVID-19 observed prior to or on the same day as this diagnosis. Hospitalisation with COVID-19 was identified as a hospital admission where the individual had a compatible COVID-19 clinical code or positive SARS-CoV-2 test over the 21 days prior to their admission up to three days after admission. ICU admission during hospitalisation was identified in a similar manner. A COVID-19 death was defined as a death where an individual had a compatible COVID-19 clinical code or positive SARS-CoV-2 test recorded in the 28 days preceding their death, with deaths identified from regional registers linked at the person-level.

To assess the impact of using alternative definitions for outpatient diagnosis of COVID-19, we explored four further definitions: PCR positive test, PCR or antigen positive test, COVID-19 diagnosis (narrow definition), and COVID-19 diagnosis (broad definition). While the broad definition for diagnoses allowed for included codes such as “ Coronavirus infection” and “ Suspected COVID-19”, the narrow definition only included codes specific to COVID-19, such as “ Disease caused by 2019 novel coronavirus”.

#### Variables

The age, as of 1^st^ March 2020 (the index date for all individuals), and sex of study participants were extracted. Using their most recent observation, individuals’ body mass index (BMI) and smoking status (classified as never smoker, former smoker, or current smoker) were also obtained. Individuals’ comorbidities and medication use was also summarised relative to the 1^st^ March 2020. The comorbidities included were autoimmune diseases, asthma, malignant neoplastic disease, diabetes mellitus, heart disease, hypertensive disorder, renal impairment, chronic obstructive lung disease [COPD], and dementia. These health conditions were identified based on an individual’s entire observed history prior to the index date. In addition, for those individuals in the outpatient diagnosis of COVID-19 cohort, symptoms recorded within two days prior to two days after index date were also identified. The following symptoms were considered: cough, dyspnea, diarrhea, headache, fever, one of anosmia, hyposmia, or dysgeusia, either malaise or fatigue, and pain.

#### Descriptive analysis

The characteristics of the study population as a whole and each of the COVID-19 outcome cohorts were summarised, with counts and percentages for categorical variables and median and interquartile ranges (IQR) for continuous variables. Cohort entry over time is plotted for the study cohorts. The proportion of persons in the outpatient COVID-19 cohort with a symptom of interest is summarised, stratified by calendar month.

All analysis code (including phenotype definitions) used in the study has been made publicly available at: https://github.com/SIDIAP/CovidCdmSummary

## Results

### Mapping to the OMOP CDM

The SIDIAP CDM contained information on 8,022,374 unique individuals. These people had 228,003,476 records in the condition occurrence table, 542,711,011 records in the drug exposure table, and 1,317,159,817 records in the measurement table. Of the 3,486 data quality checks run against the database, 3,456 passed (99%). The remaining 30 checks that failed were considered not to be pertinent, and each of these is summarised in Supplementary Table 1.

### COVID-19 outcomes

A total of 5,870,274 individuals were included in the general population cohort of people alive in the database as of 1^st^ March 2020. Over observed follow-up, 604,472 had either an outpatient COVID-19 diagnosis or positive test result, 58,991 were hospitalised with COVID-19, 5,642 had an ICU admission with COVID-19, and 11,233 had a COVID-19 death.

The distribution of age for each study cohort, stratified by sex, is shown in Figure 1. The average age of the general population study cohort was 43 years (IQR: 25 to 59), with 50.7% female. Median age was higher among outcome cohorts, most notably among those with a COVID-19 death who had an average age of 85 (78 to 90). Patients admitted to ICU, however, were younger than those admitted to hospital in general (63 [53 to 71] compared to 65 [51 to 78]). While the outpatient COVID-19 cohort was majority female (54%), those hospitalised were more typically male (55%), and those with a COVID-19 death were close to equally distributed by sex (49.1% were female). Patients admitted to ICU were though majority male (67%). Comorbidities were generally more common among those with a COVID-19 outcome compared to the general population, see Table 2. For example, prevalence of diabetes and hypertension were 23% and 45% among those hospitalised with COVID-19 which compared to 7% and 17% in the general population.

**Table 2.** Characteristics of the SIDIAP COVID-19 study cohorts

|  | General population | Outpatient COVID-19 diagnosis or positive test | Hospitalised with COVID-19 | ICU admission with COVID-19 | Died with COVID-19 |
| --- | --- | --- | --- | --- | --- |
| N | 5,870,274 | 604,472 | 58,991 | 5,642 | 11,233 |
| Age (median [IQR]) | 43 [25 to 59] | 41 [25 to 55] | 65 [51 to 78] | 63 [53 to 71] | 85 [78 to 90] |
| Age group (n (%)) |  |  |  |  |  |
| Under 20 | 1,158,540 (19.7%) | 111,037 (18.4%) | 644 (1.1%) | 39 (0.7%) | <5 |
| 20 to 29 | 629,785 (10.7%) | 80,548 (13.3%) | 1,888 (3.2%) | 89 (1.6%) | 6 (0.1%) |
| 30 to 39 | 798,397 (13.6%) | 91,377 (15.1%) | 3,654 (6.2%) | 277 (4.9%) | 22 (0.2%) |
| 40 to 49 | 1,010,564 (17.2%) | 113,666 (18.8%) | 7,023 (11.9%) | 651 (11.5%) | 76 (0.7%) |
| 50 to 59 | 824,798 (14.1%) | 87,726 (14.5%) | 10,053 (17.0%) | 1,204 (21.3%) | 270 (2.4%) |
| 60 to 69 | 626,418 (10.7%) | 50,865 (8.4%) | 10,655 (18.1%) | 1,691 (30.0%) | 781 (7.0%) |
| 70 to 79 | 477,436 (8.1%) | 33,403 (5.5%) | 11,822 (20.0%) | 1,416 (25.1%) | 2,287 (20.4%) |
| 80 or older | 344,336 (5.9%) | 35,850 (5.9%) | 13,252 (22.5%) | 275 (4.9%) | 7,788 (69.3%) |
| Sex: Male (n (%)) | 2,896,281 (49.3%) | 280,092 (46.3%) | 32,174 (54.5%) | 3,790 (67.2%) | 5,712 (50.9%) |
| Years of prior observation time (median [IQR]) | 14.2 [11.9 to 14.2] | 14.2 [12.7 to 14.2] | 14.2 [14.2 to 14.2] | 14.2 [14.2 to 14.2] | 14.2 [14.2 to 14.2] |
| Smoking status (n (%)) |  |  |  |  |  |
| Current smoker | 739,900 (12.6%) | 64,469 (10.7%) | 3,595 (6.1%) | 320 (5.7%) | 492 (4.4%) |
| Ex-smoker | 789,023 (13.4%) | 87,556 (14.5%) | 16,547 (28.1%) | 1,904 (33.7%) | 3,601 (32.1%) |
| Non-smoker | 2,111,283 (36.0%) | 252,115 (41.7%) | 24,906 (42.2%) | 2,246 (39.8%) | 4,158 (37.0%) |
| Missing | 2,230,068 (38.0%) | 200,332 (33.1%) | 13,943 (23.6%) | 1,172 (20.8%) | 2,982 (26.5%) |
| Years since smoking status observation (median [IQR]) | 7.4 [3.6 to 11.0] | 7.3 [3.6 to 10.8] | 8.5 [4.4 to 11.8] | 8.0 [4.2 to 11.4] | 9.6 [5.5 to 12.4] |
| BMI (n (%)) | 26 [22 to 29] | 26 [22 to 29] | 29 [26 to 32] | 30 [27 to 34] | 28 [25 to 31] |
| Years since BMI observation (median [IQR]) | 1.8 [0.6 to 4.4] | 1.9 [0.6 to 4.6] | 1.0 [0.3 to 2.6] | 0.9 [0.3 to 2.3] | 1.0 [0.3 to 2.4] |
| Comorbidities (n (%)) |  |  |  |  |  |
| Autoimmune disease | 86,516 (1.5%) | 9,386 (1.6%) | 1,890 (3.2%) | 168 (3.0%) | 478 (4.3%) |
| Asthma | 306,554 (5.2%) | 37,323 (6.2%) | 4,054 (6.9%) | 327 (5.8%) | 661 (5.9%) |
| Malignant neoplastic disease | 350,077 (6.0%) | 32,408 (5.4%) | 9,810 (16.6%) | 764 (13.5%) | 3,255 (29.0%) |
| Diabetes mellitus | 430,519 (7.3%) | 44,746 (7.4%) | 13,649 (23.1%) | 1,406 (24.9%) | 3,793 (33.8%) |
| Heart disease | 615,536 (10.5%) | 61,661 (10.2%) | 18,569 (31.5%) | 1,491 (26.4%) | 6,482 (57.7%) |
| Hypertensive disorder | 982,299 (16.7%) | 96,618 (16.0%) | 26,234 (44.5%) | 2,482 (44.0%) | 7,449 (66.3%) |
| Renal impairment | 259,620 (4.4%) | 26,289 (4.3%) | 11,032 (18.7%) | 800 (14.2%) | 4,792 (42.7%) |
| COPD | 165,181 (2.8%) | 15,578 (2.6%) | 6,328 (10.7%) | 536 (9.5%) | 2,076 (18.5%) |
| Dementia | 67,172 (1.1%) | 13,431 (2.2%) | 3,388 (5.7%) | 47 (0.8%) | 3,173 (28.2%) |

IQR: interquartile range; BMI: body mass index; COPD: chronic obstructive lung disease; ICU: intensive care unit

**Figure 1.**
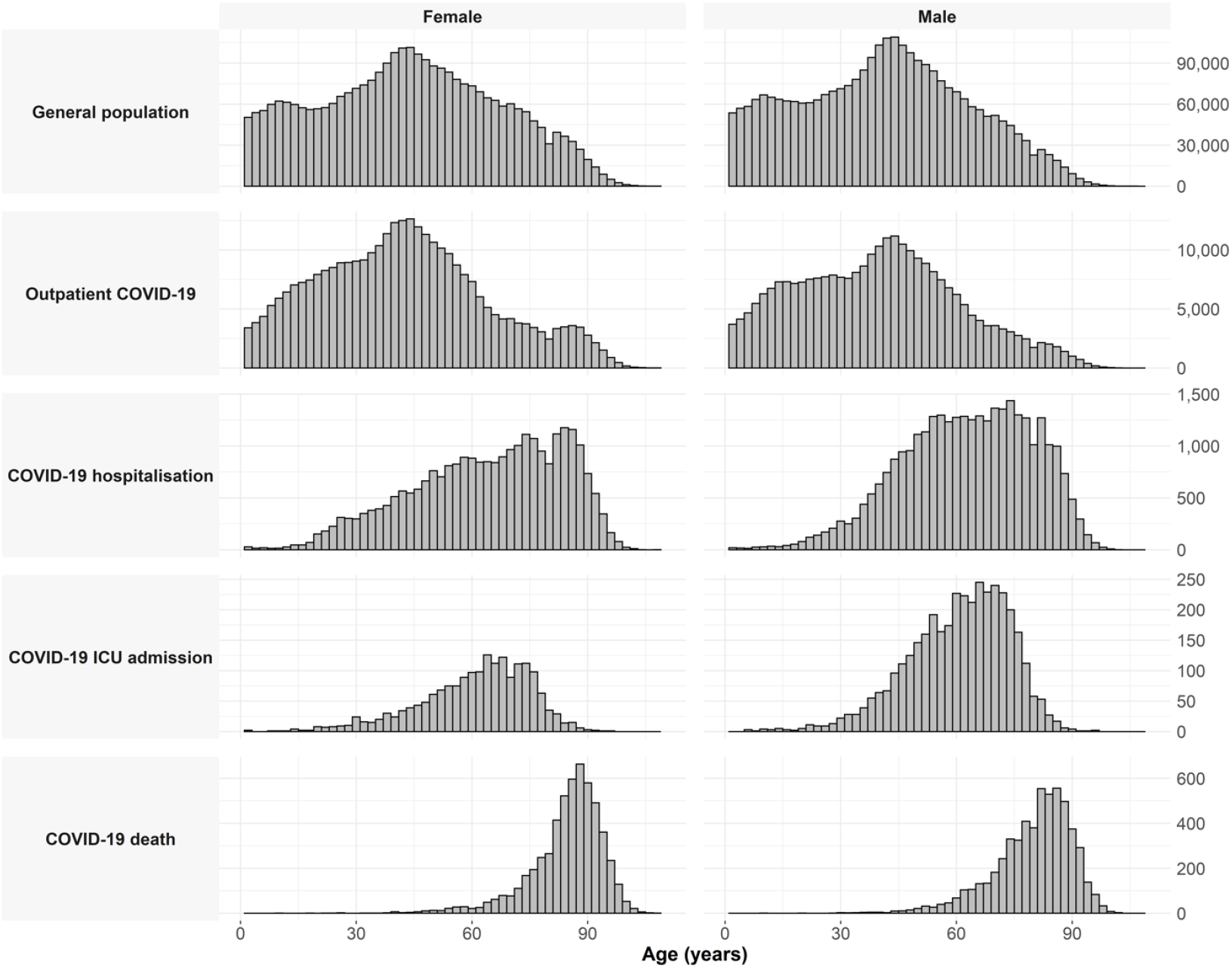
Histogram of age, stratified by sex, for the general population and each COVID-19 outcome cohort

Cohort entry over calendar time, stratified by age, is shown in Figure 2. The various waves of COVID-19 can be seen, along with the much greater number of cases of COVID-19 hospitalisations, ICU admission, and deaths among the older age groups. The highest number of outpatient COVID-19 cases did though occur among the younger age group.

**Figure 2.**
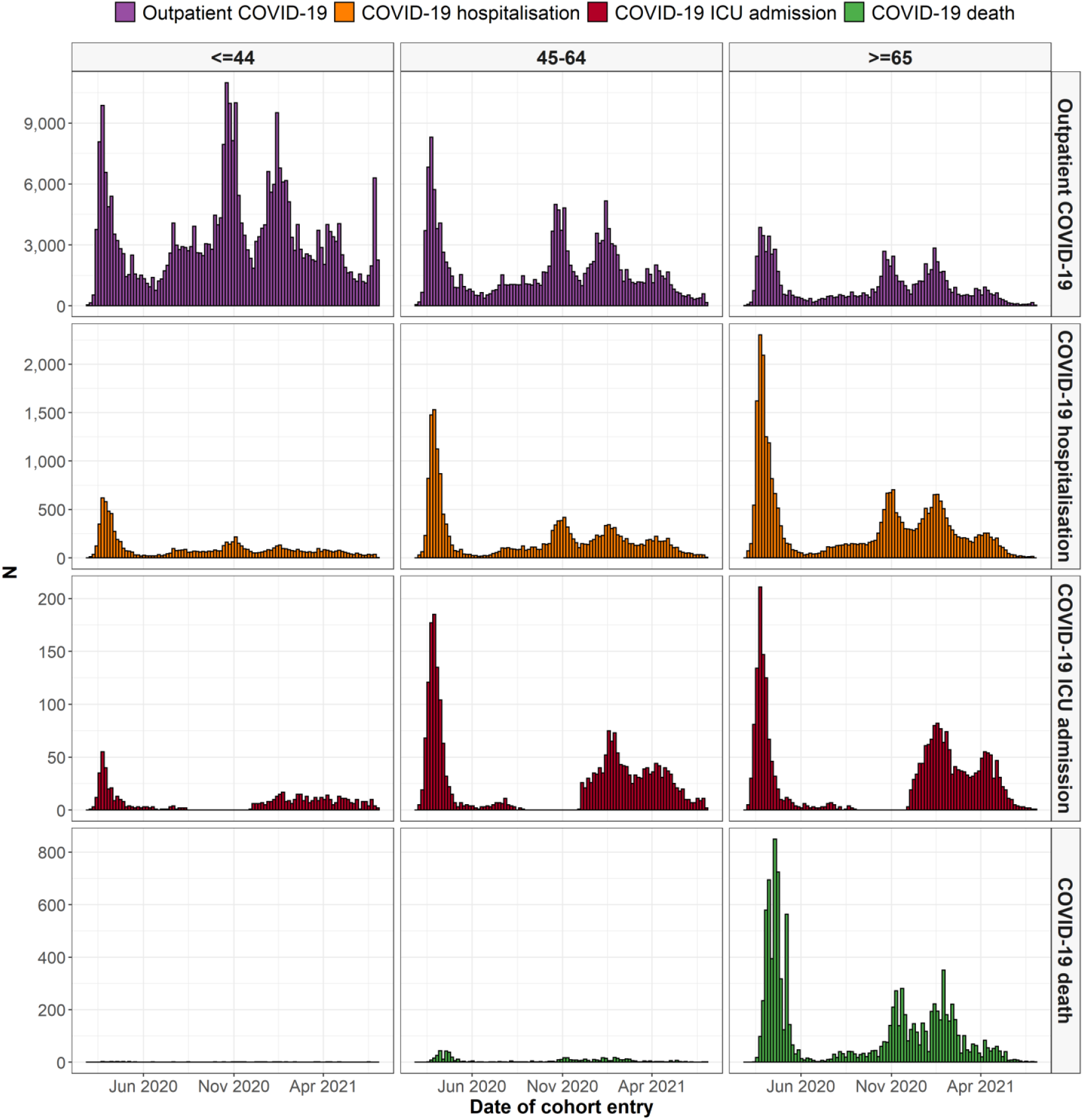
COVID-19 outcome cohort entry over calendar time, stratified by age group

Capture of COVID-19 symptoms over calendar time is shown in Figure 3 and stratified by age group in Supplementary Figure 3 and stratified by sex in Supplementary Figure 4. Cough and fever were the most common symptoms identified, but all symptoms had a prevalence of less than 15% and with substantial changes over time.

**Figure 3.**
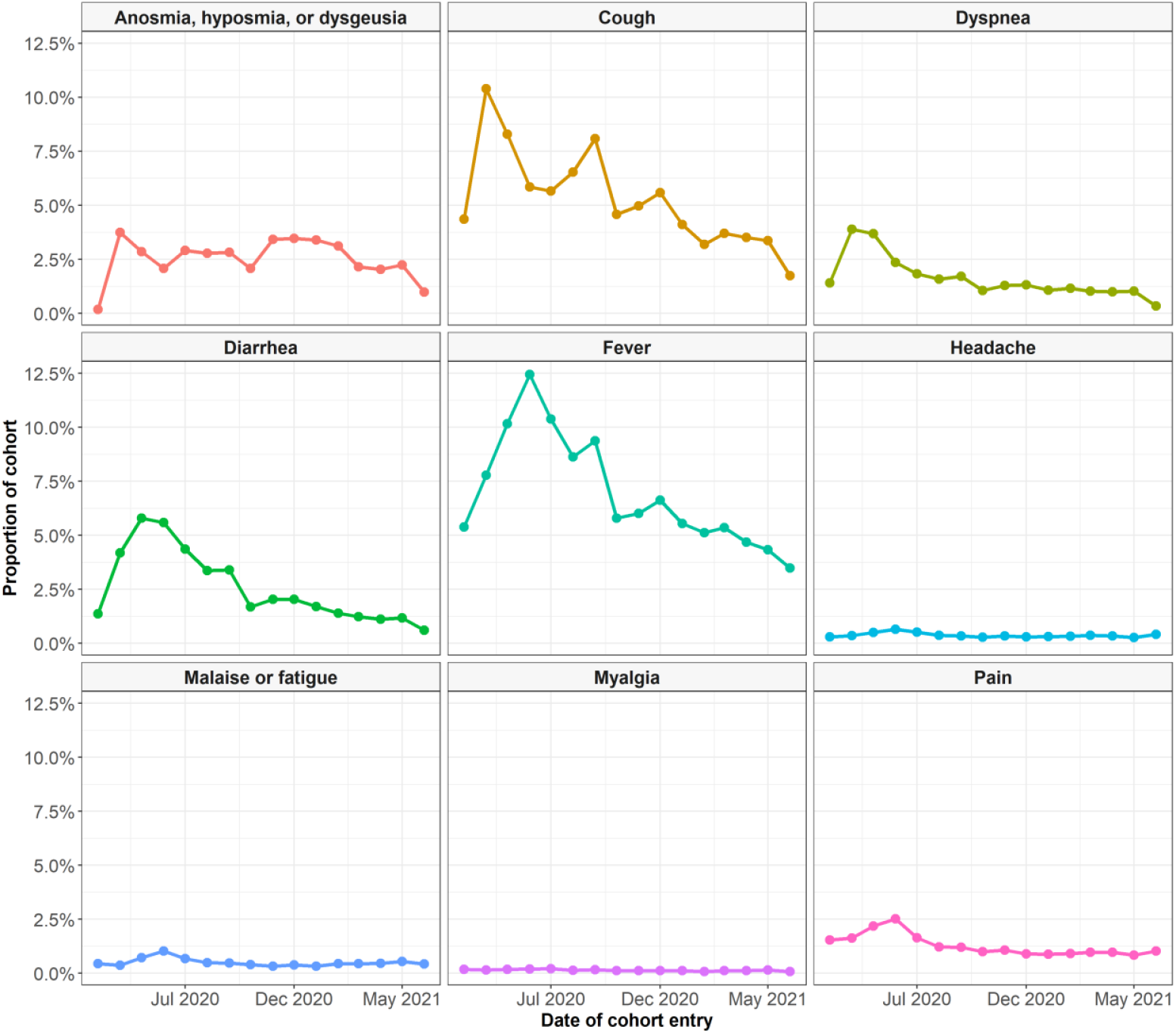
Symptoms recorded at time of outpatient COVID-19 diagnosis or positive test

The impact of different definitions for outpatient COVID-19 is shown in Figure 4, where cohort entry over calendar time is depicted. While from September 2020 definitions were generally in accordance, the first wave of COVID-19 in Catalonia was only identified when including the broad COVID-19 diagnosis definition.

**Figure 4.**
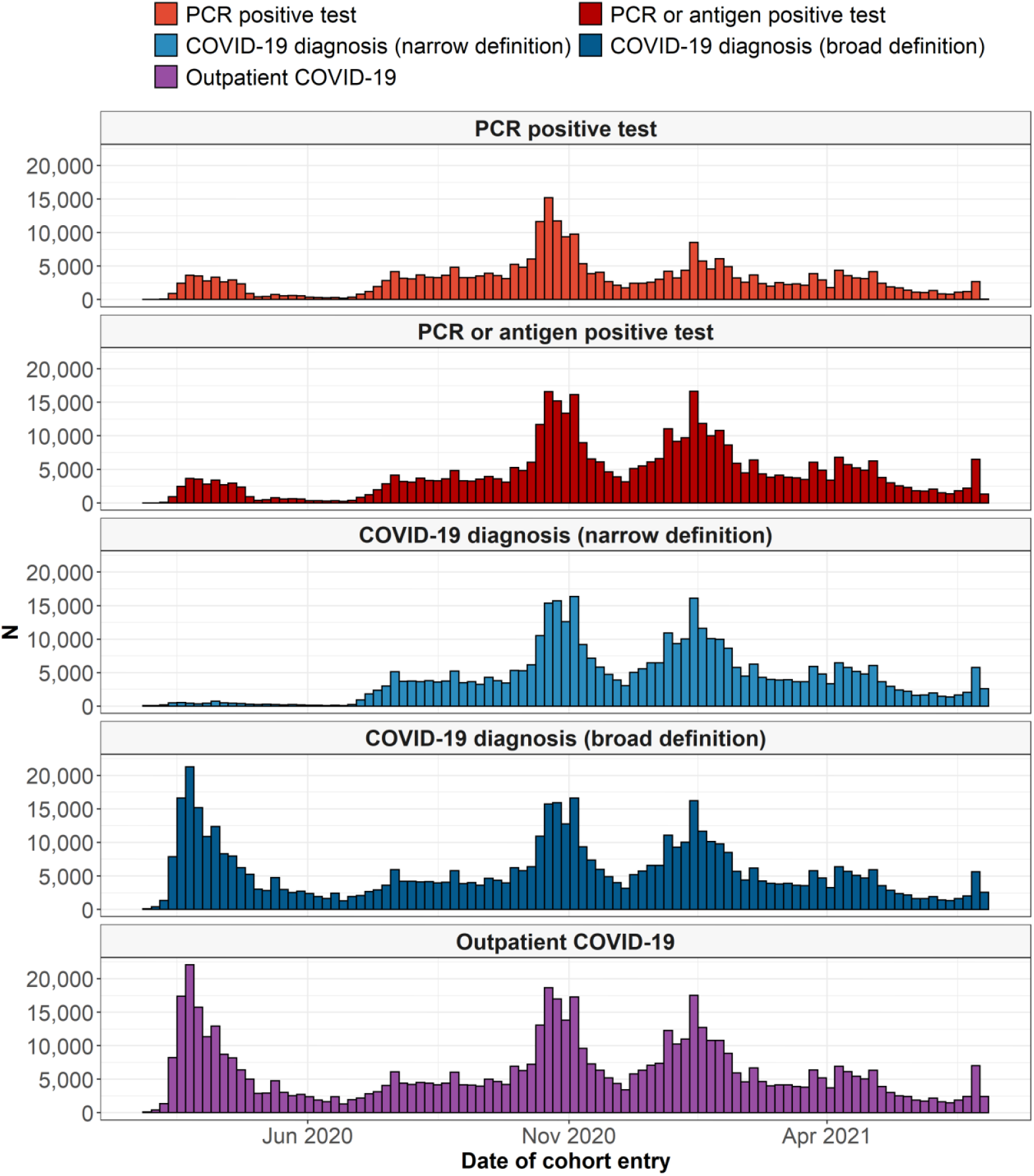
Outpatient COVID-19 cohort entry over calendar time

## Discussion

In this study we have described the development of a wide-ranging dataset for COVID-19. With more than 3,000 data quality checks performed to assess data quality, the resulting database can be considered fit for use to inform appropriate research questions. Moreover, the use of the OMOP CDM will facilitate its use for both single database studies and distributed network research. Demonstrating the breadth of data captured, a descriptive analysis of various COVID-19 outcomes among the general population has been performed, providing a broad overview of COVID-19 in Catalonia and the characteristics of the individuals affected.

Numerous health care databases, including both claims and electronic health care data, have previously been mapped to the OMOP CDM.^13,14^ The European Health Data and Evidence Network (EHDEN, www.ehden.eu) is building a large, federated network of data sources mapped to the OMOP CDM and SIDIAP is an EHDEN data partner. The mapping of SIDIAP data to the OMOP CDM has already facilitated its use in several international network studies on COVID-19. Examples include a characterisation study of patients hospitalised with COVID-19,^15^ a study to develop a prediction model for hospitalisation and mortality among patients diagnosed with COVID-19,^16^ and another which assessed the impact of angiotensin converting enzyme inhibitors and angiotensin receptor blockers on the risk of COVID-19.^17^ In other studies based solely on SIDIAP data mapped to the OMOP CDM, multi-state modelling was used to describe COVID-19 patient trajectories in Catalonia and the impact of cancer and obesity on these trajectories.^18–20^ The impact of the pandemic on trends in diagnoses of anxiety and depression has also been studied.^21^

In our descriptive analysis we found that individuals with a COVID-19 outcome were typically older and had more comorbidities than the general population. This was particularly pronounced for the most severe outcomes studied. This is in concordance with research to date, with numerous studies finding older age to be associated with worse outcomes in COVID-19.^22–25^ While those with an outpatient COVID-19 diagnosis or positive test were more often female in our data, those hospitalised were majority male, as were 67% of those admitted to ICU. People who died with COVID-19 were almost equally distributed by sex. Previous research studies have reported mixed results for diagnoses and positive tests, for example two studies from the UK which reported a higher risk of testing positive for SARS-CoV-2 among men,^26,27^ while a study from China found there to have been a higher attack rate among women.^28^ A range of studies have though previously found males to be at an increased risk of severest outcomes.^23,24,29^

The importance of appropriate phenotyping when using routinely collected data is also demonstrated when comparing alternative definitions of an outpatient COVID-19 case in our data. A definition that relied solely on testing for SARS-CoV-2 or using a narrow set of diagnosis codes would have missed many of the COVID-19 cases from the first wave, a time when testing was not widely available and medical vocabularies had not yet introduced COVID-19 specific codes.

### Strengths and limitations

Much of the COVID-19 literature is based on studies where study populations have been drawn from people hospitalised with COVID-19, tested for infection, or who volunteered to participate in a study. Such studies can be subject to a number of biases, in particular collider bias which can lead to the reporting associations that do not exist for the general population or by attenuating, inflating or reversing the sign of true associations.^30^ This underscores the importance of developing comprehensive datasets to generate the reliable evidence required to inform decision-making related to the pandemic. With more than half a million outpatient cases of COVID-19 captured and a breadth of data capture that allows for comparisons with the general population and subsequent hospital care to be described, the mapped SIDIAP database described here is one such resource.

While electronic health record data brings numerous opportunities, with the data collected for non-research purposes careful curation is required. Using a well-established common data model, meant that existing open-source tools could be used to evaluate data quality and that research studies can be run in a distributed manner. This has allowed the database to already have been used in a number of international network research studies, with standardised analytic packages and only aggregated results sets shared.

One limitation of the dataset has been seen with the likely underreporting of COVID-19 symptoms. The estimates drawn from this database are much lower than reported in studies informed by self-reported patient data.^31,32^ Capture of symptoms would likely be improved if free text data recorded during primary care visits was also mapped to the OMOP CDM. However, underreporting of symptoms also likely reflects the nature of electronic health care records not designed for specific research questions. Other limitations include lack of hospital prescribing of medicines and lab data, while SARS-CoV-2 variants and contact tracing are also not captured. Vaccination records are available, and linkage to these ongoing.

## Conclusion

We have established a wide-ranging COVID-19 dataset that captures COVID-19 diagnoses, SARS-CoV-2 test results, hospitalisations, and deaths in Catalonia, Spain. In this study we have summarised the creation of this dataset and described observed COVID-19 outcomes and summarised the characteristics of those individuals affected.

## Data Availability

In accordance with current European and national law, the data used in this study is only available for the researchers participating in this study. Thus, we are not allowed to distribute or make publicly available the data to other parties. However, researchers from public institutions can request data from SIDIAP if they comply with certain requirements. Further information is available online (https://www.sidiap.org/index.php/menu-solicitudesen/application-proccedure) or by contacting SIDIAP.

## Acknowledgements

We would like to acknowledge the patients who suffered or died from this devastating disease, and these patient’s families and carers. We would also like to thank the healthcare professionals in the Catalan healthcare system involved in the management of COVID-19 during these challenging times, from primary care to intensive care units.

## Ethics approval

This study was approved by the Clinical Research Ethics Committee of the IDIAPJGol (project code: 20/070-PCV).

## Appendix

**Supplementary Table 1.**
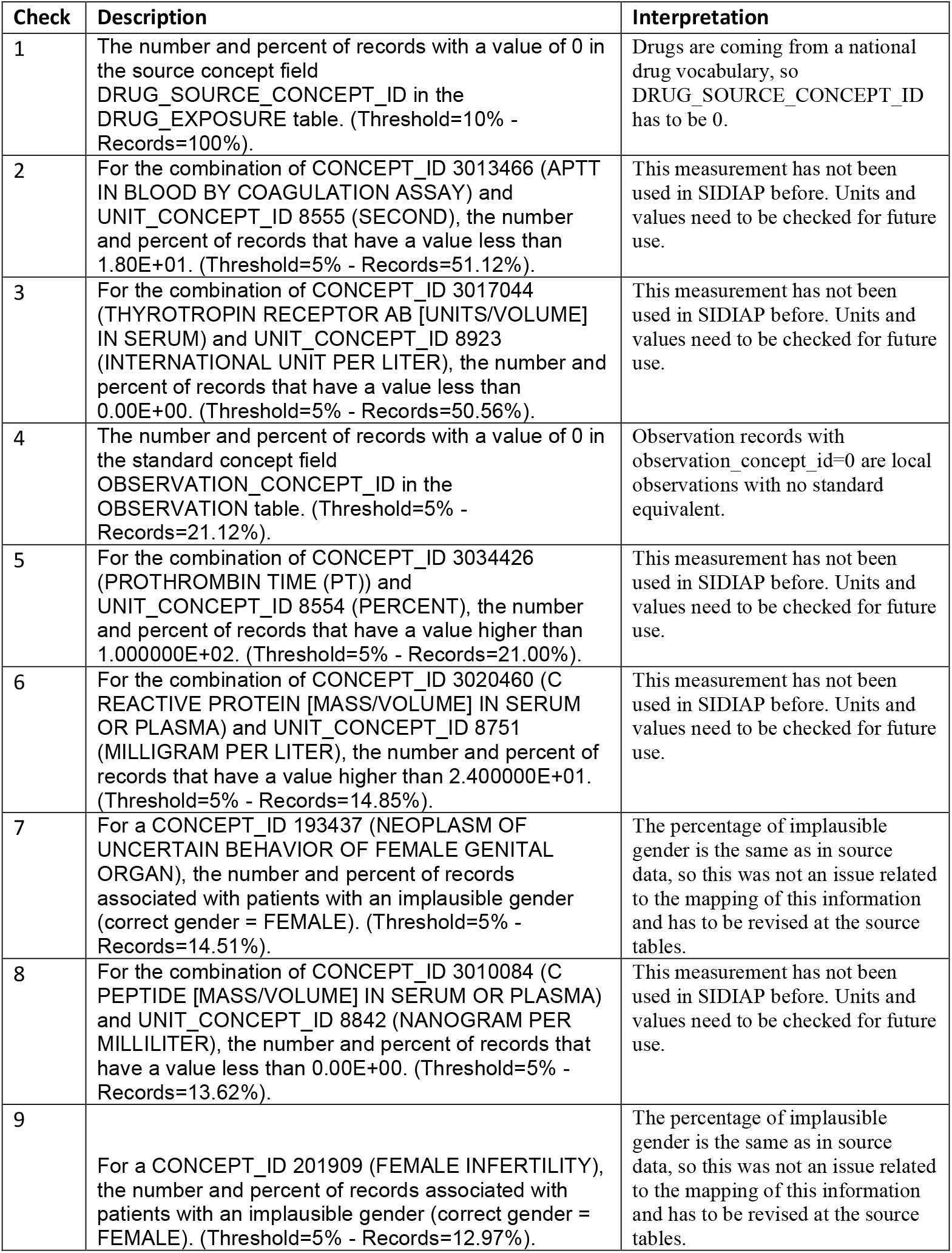
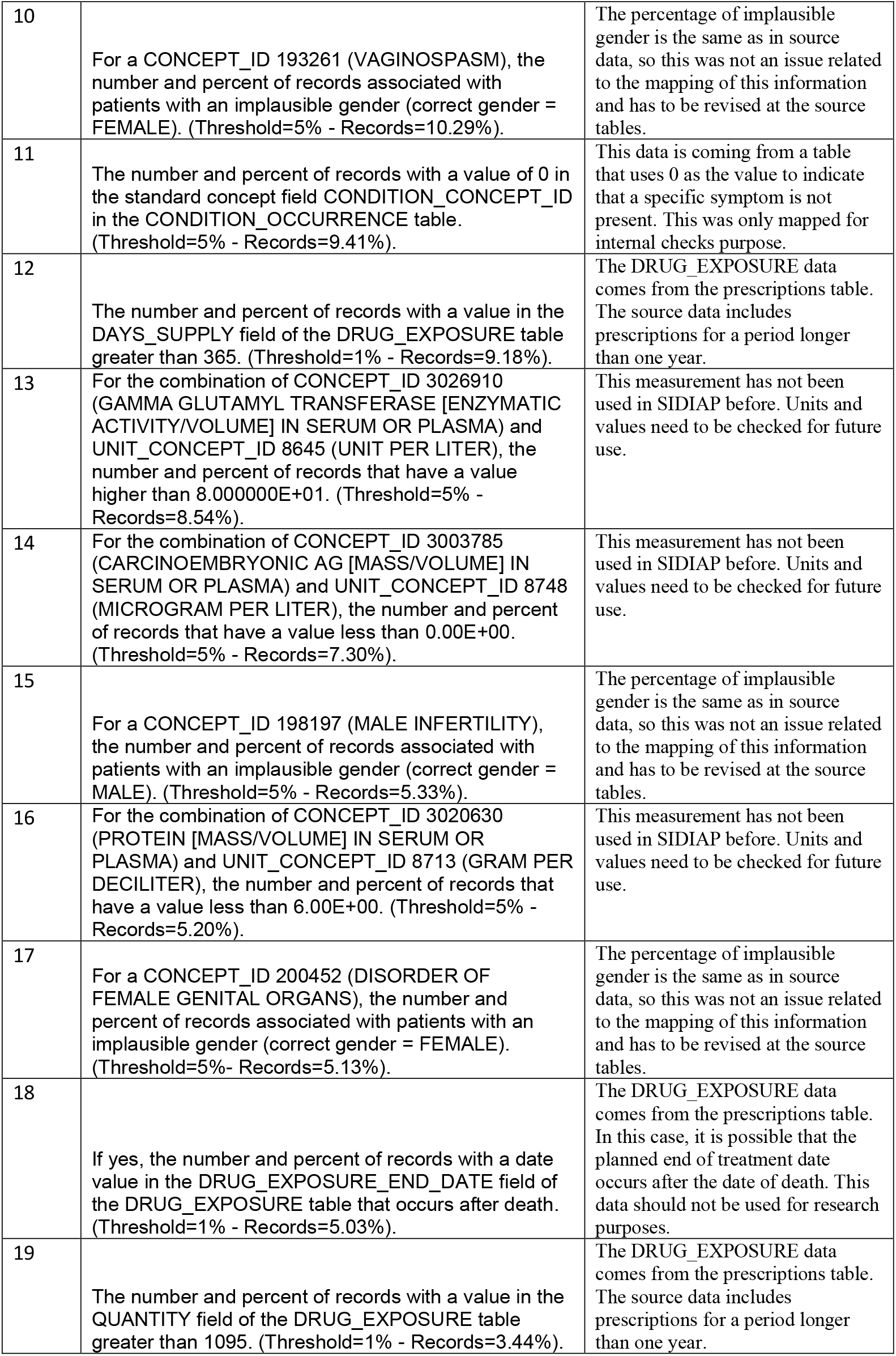
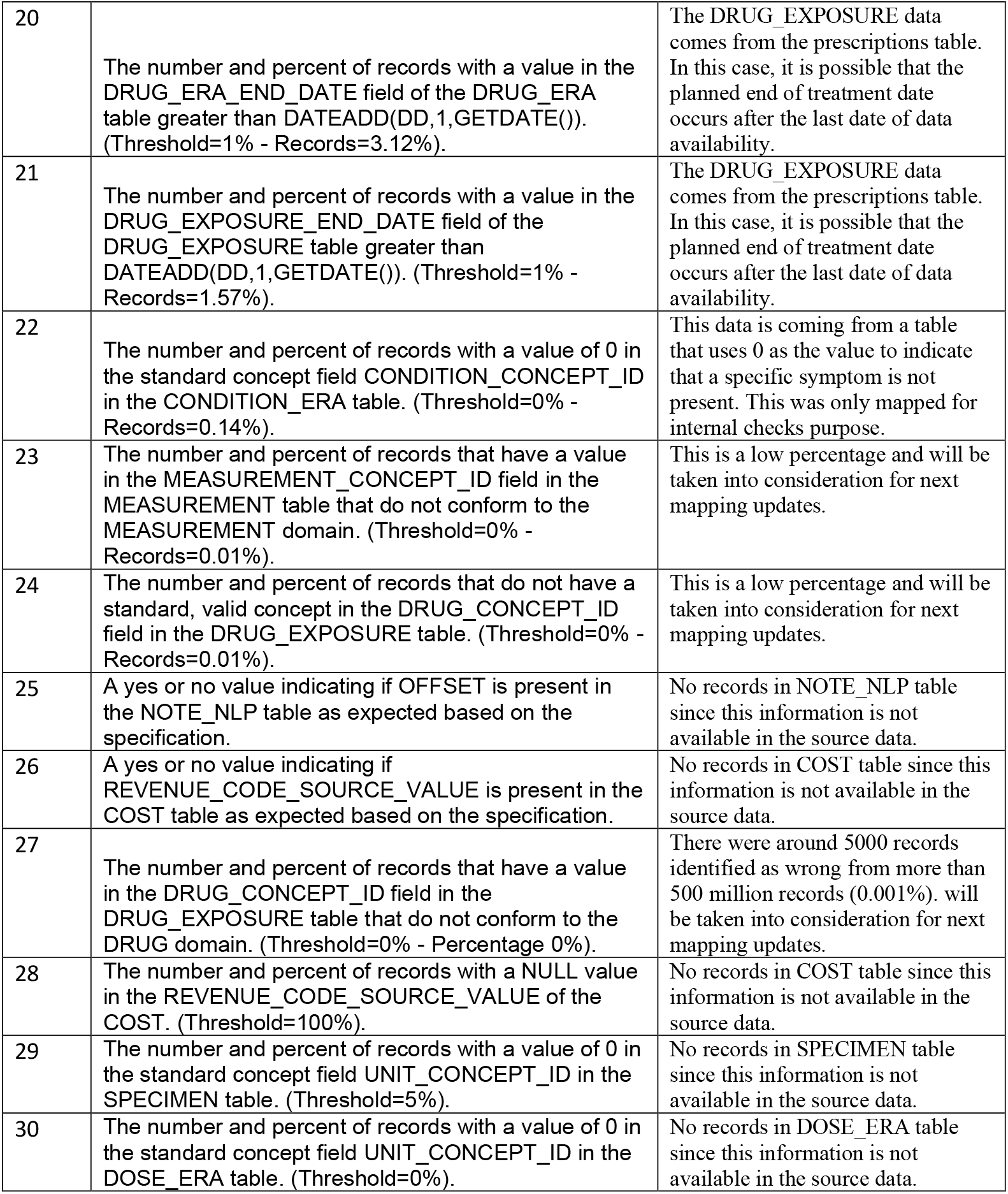
Failed data quality checks

| Check | Description | Interpretation |
| --- | --- | --- |
| 1 | The number and percent of records with a value of 0 in the source concept field DRUG_SOURCE_CONCEPT_ID in the DRUG_EXPOSURE table. (Threshold=10% - Records=100%). | Drugs are coming from a national drug vocabulary, so DRUG_SOURCE_CONCEPT_ID has to be 0. |
| 2 | For the combination of CONCEPT_ID 3013466 (APTT IN BLOOD BY COAGULATION ASSAY) and UNIT_CONCEPT_ID 8555 (SECOND), the number and percent of records that have a value less than 1.80E+01. (Threshold=5% - Records=51.12%). | This measurement has not been used in SIDIAP before. Units and values need to be checked for future use. |
| 3 | For the combination of CONCEPT_ID 3017044 (THYROTROPIN RECEPTOR AB [UNITS/VOLUME] IN SERUM) and UNIT_CONCEPT_ID 8923 (INTERNATIONAL UNIT PER LITER), the number and percent of records that have a value less than 0.00E+00. (Threshold=5% - Records=50.56%). | This measurement has not been used in SIDIAP before. Units and values need to be checked for future use. |
| 4 | The number and percent of records with a value of 0 in the standard concept field OBSERVATION_CONCEPT_ID in the OBSERVATION table. (Threshold=5% - Records=21.12%). | Observation records with observation_concept_id=0 are local observations with no standard equivalent. |
| 5 | For the combination of CONCEPT_ID 3034426 (PROTHROMBIN TIME (PT)) and UNIT_CONCEPT_ID 8554 (PERCENT), the number and percent of records that have a value higher than 1.000000E+02. (Threshold=5% - Records=21.00%). | This measurement has not been used in SIDIAP before. Units and values need to be checked for future use. |
| 6 | For the combination of CONCEPT_ID 3020460 (C REACTIVE PROTEIN [MASS/VOLUME] IN SERUM OR PLASMA) and UNIT_CONCEPT_ID 8751 (MILLIGRAM PER LITER), the number and percent of records that have a value higher than 2.400000E+01. (Threshold=5% - Records=14.85%). | This measurement has not been used in SIDIAP before. Units and values need to be checked for future use. |
| 7 | For a CONCEPT_ID 193437 (NEOPLASM OF UNCERTAIN BEHAVIOR OF FEMALE GENITAL ORGAN), the number and percent of records associated with patients with an implausible gender (correct gender = FEMALE). (Threshold=5% - Records=14.51%). | The percentage of implausible gender is the same as in source data, so this was not an issue related to the mapping of this information and has to be revised at the source tables. |
| 8 | For the combination of CONCEPT_ID 3010084 (C PEPTIDE [MASS/VOLUME] IN SERUM OR PLASMA) and UNIT_CONCEPT_ID 8842 (NANOGRAM PER MILLILITER), the number and percent of records that have a value less than 0.00E+00. (Threshold=5% - Records=13.62%). | This measurement has not been used in SIDIAP before. Units and values need to be checked for future use. |
| 9 | For a CONCEPT_ID 201909 (FEMALE INFERTILITY), the number and percent of records associated with patients with an implausible gender (correct gender = FEMALE). (Threshold=5% - Records=12.97%). | The percentage of implausible gender is the same as in source data, so this was not an issue related to the mapping of this information and has to be revised at the source tables. |
| 10 | For a CONCEPT_ID 193261 (VAGINOSPASM), the number and percent of records associated with patients with an implausible gender (correct gender = FEMALE). (Threshold=5% - Records=10.29%). | The percentage of implausible gender is the same as in source data, so this was not an issue related to the mapping of this information and has to be revised at the source tables. |
| 11 | The number and percent of records with a value of 0 in the standard concept field CONDITION_CONCEPT_ID in the CONDITION_OCCURRENCE table. (Threshold=5% - Records=9.41%). | This data is coming from a table that uses 0 as the value to indicate that a specific symptom is not present. This was only mapped for internal checks purpose. |
| 12 | The number and percent of records with a value in the DAYS_SUPPLY field of the DRUG_EXPOSURE table greater than 365. (Threshold=1% - Records=9.18%). | The DRUG_EXPOSURE data comes from the prescriptions table. The source data includes prescriptions for a period longer than one year. |
| 13 | For the combination of CONCEPT_ID 3026910 (GAMMA GLUTAMYL TRANSFERASE [ENZYMATIC ACTIVITY/VOLUME] IN SERUM OR PLASMA) and UNIT_CONCEPT_ID 8645 (UNIT PER LITER), the number and percent of records that have a value higher than 8.000000E+01. (Threshold=5% - Records=8.54%). | This measurement has not been used in SIDIAP before. Units and values need to be checked for future use. |
| 14 | For the combination of CONCEPT_ID 3003785 (CARCINOEMBRYONIC AG [MASS/VOLUME] IN SERUM OR PLASMA) and UNIT_CONCEPT_ID 8748 (MICROGRAM PER LITER), the number and percent of records that have a value less than 0.00E+00. (Threshold=5% - Records=7.30%). | This measurement has not been used in SIDIAP before. Units and values need to be checked for future use. |
| 15 | For a CONCEPT_ID 198197 (MALE INFERTILITY), the number and percent of records associated with patients with an implausible gender (correct gender = MALE). (Threshold=5% - Records=5.33%). | The percentage of implausible gender is the same as in source data, so this was not an issue related to the mapping of this information and has to be revised at the source tables. |
| 16 | For the combination of CONCEPT_ID 3020630 (PROTEIN [MASS/VOLUME] IN SERUM OR PLASMA) and UNIT_CONCEPT_ID 8713 (GRAM PER DECILITER), the number and percent of records that have a value less than 6.00E+00. (Threshold=5% - Records=5.20%). | This measurement has not been used in SIDIAP before. Units and values need to be checked for future use. |
| 17 | For a CONCEPT_ID 200452 (DISORDER OF FEMALE GENITAL ORGANS), the number and percent of records associated with patients with an implausible gender (correct gender = FEMALE). (Threshold=5%- Records=5.13%). | The percentage of implausible gender is the same as in source data, so this was not an issue related to the mapping of this information and has to be revised at the source tables. |
| 18 | If yes, the number and percent of records with a date value in the DRUG_EXPOSURE_END_DATE field of the DRUG_EXPOSURE table that occurs after death. (Threshold=1% - Records=5.03%). | The DRUG_EXPOSURE data comes from the prescriptions table. In this case, it is possible that the planned end of treatment date occurs after the date of death. This data should not be used for research purposes. |
| 19 | The number and percent of records with a value in the QUANTITY field of the DRUG_EXPOSURE table greater than 1095. (Threshold=1% - Records=3.44%). | The DRUG_EXPOSURE data comes from the prescriptions table. The source data includes prescriptions for a period longer than one year. |
| 20 | The number and percent of records with a value in the DRUG_ERA_END_DATE field of the DRUG_ERA table greater than DATEADD(DD,1,GETDATE()). (Threshold=1% - Records=3.12%). | The DRUG_EXPOSURE data comes from the prescriptions table. In this case, it is possible that the planned end of treatment date occurs after the last date of data availability. |
| 21 | The number and percent of records with a value in the DRUG_EXPOSURE_END_DATE field of the DRUG_EXPOSURE table greater than DATEADD(DD,1,GETDATE()). (Threshold=1% - Records=1.57%). | The DRUG_EXPOSURE data comes from the prescriptions table. In this case, it is possible that the planned end of treatment date occurs after the last date of data availability. |
| 22 | The number and percent of records with a value of 0 in the standard concept field CONDITION_CONCEPT_ID in the CONDITION_ERA table. (Threshold=0% - Records=0.14%). | This data is coming from a table that uses 0 as the value to indicate that a specific symptom is not present. This was only mapped for internal checks purpose. |
| 23 | The number and percent of records that have a value in the MEASUREMENT_CONCEPT_ID field in the MEASUREMENT table that do not conform to the MEASUREMENT domain. (Threshold=0% - Records=0.01%). | This is a low percentage and will be taken into consideration for next mapping updates. |
| 24 | The number and percent of records that do not have a standard, valid concept in the DRUG_CONCEPT_ID field in the DRUG_EXPOSURE table. (Threshold=0% - Records=0.01%). | This is a low percentage and will be taken into consideration for next mapping updates. |
| 25 | A yes or no value indicating if OFFSET is present in the NOTE_NLP table as expected based on the specification. | No records in NOTE_NLP table since this information is not available in the source data. |
| 26 | A yes or no value indicating if REVENUE_CODE_SOURCE_VALUE is present in the COST table as expected based on the specification. | No records in COST table since this information is not available in the source data. |
| 27 | The number and percent of records that have a value in the DRUG_CONCEPT_ID field in the DRUG_EXPOSURE table that do not conform to the DRUG domain. (Threshold=0% - Percentage 0%). | There were around 5000 records identified as wrong from more than 500 million records (0.001%). will be taken into consideration for next mapping updates. |
| 28 | The number and percent of records with a NULL value in the REVENUE_CODE_SOURCE_VALUE of the COST. (Threshold=100%). | No records in COST table since this information is not available in the source data. |
| 29 | The number and percent of records with a value of 0 in the standard concept field UNIT_CONCEPT_ID in the SPECIMEN table. (Threshold=5%). | No records in SPECIMEN table since this information is not available in the source data. |
| 30 | The number and percent of records with a value of 0 in the standard concept field UNIT_CONCEPT_ID in the DOSE_ERA table. (Threshold=0%). | No records in DOSE_ERA table since this information is not available in the source data. |

**Supplementary Figure 1.**
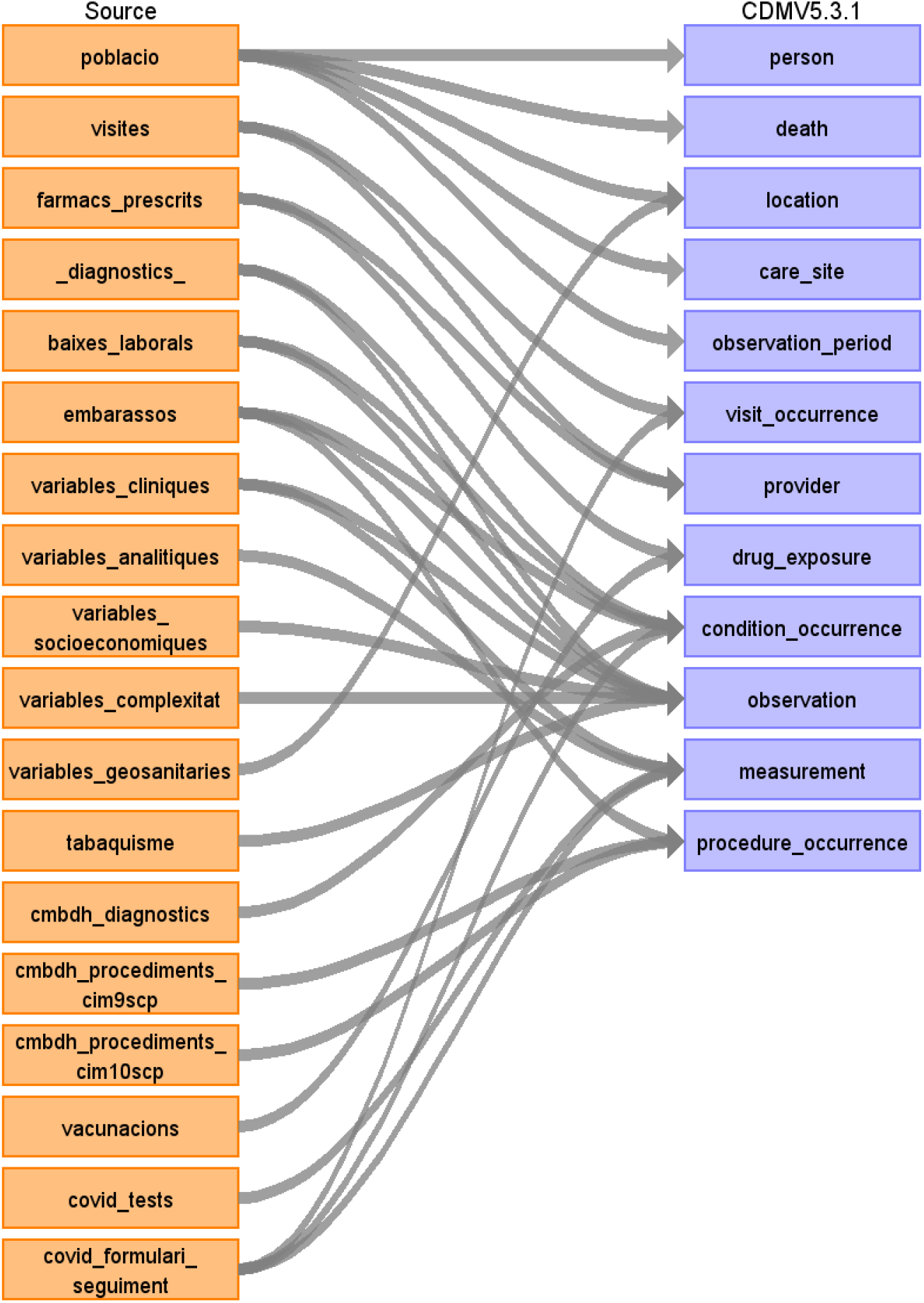
Source tables to OMOP-CDM tables conversion diagram

**Supplementary Figure 2.**
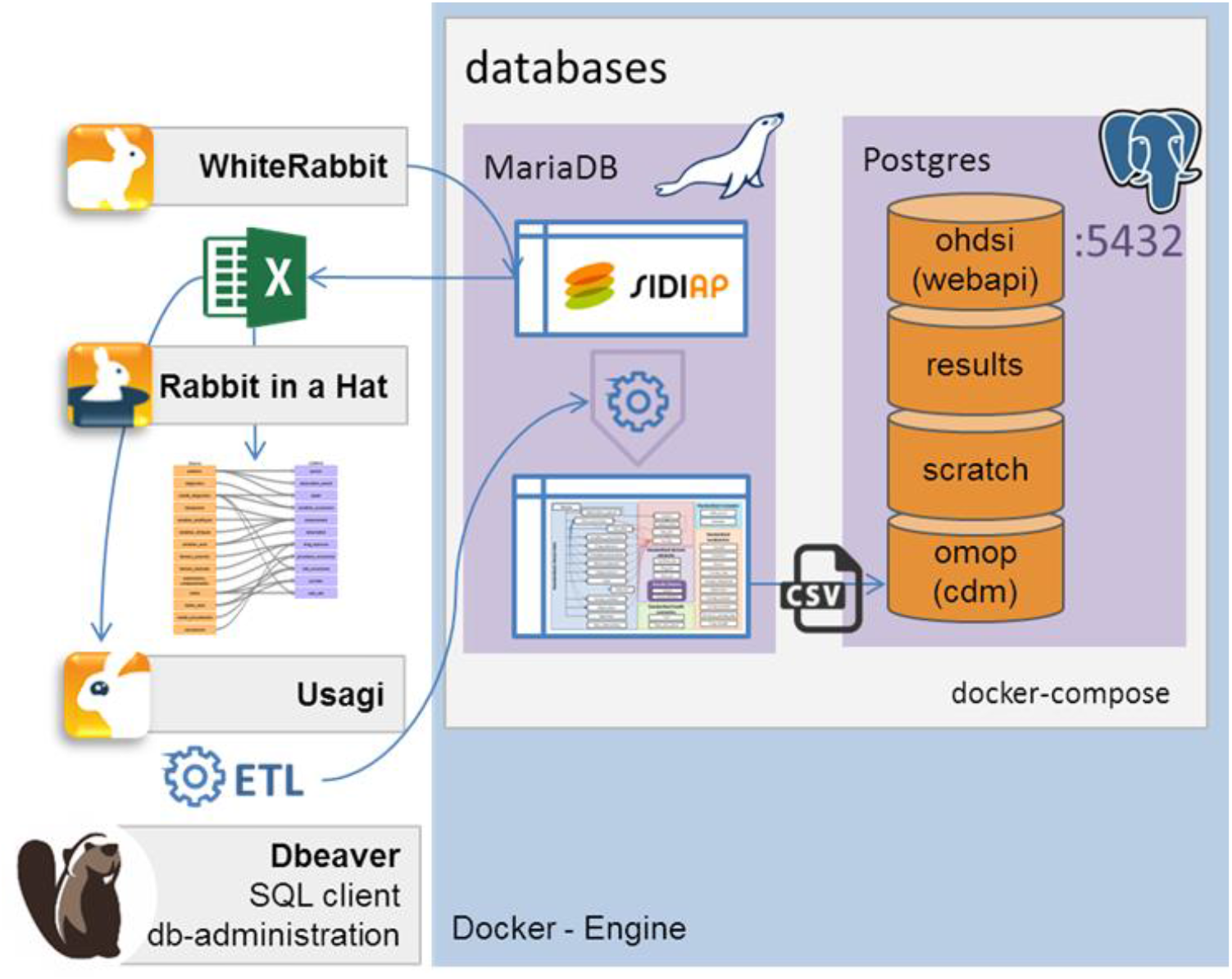
Extract, transform, and load (ETL) framework

**Supplementary Figure 3.**
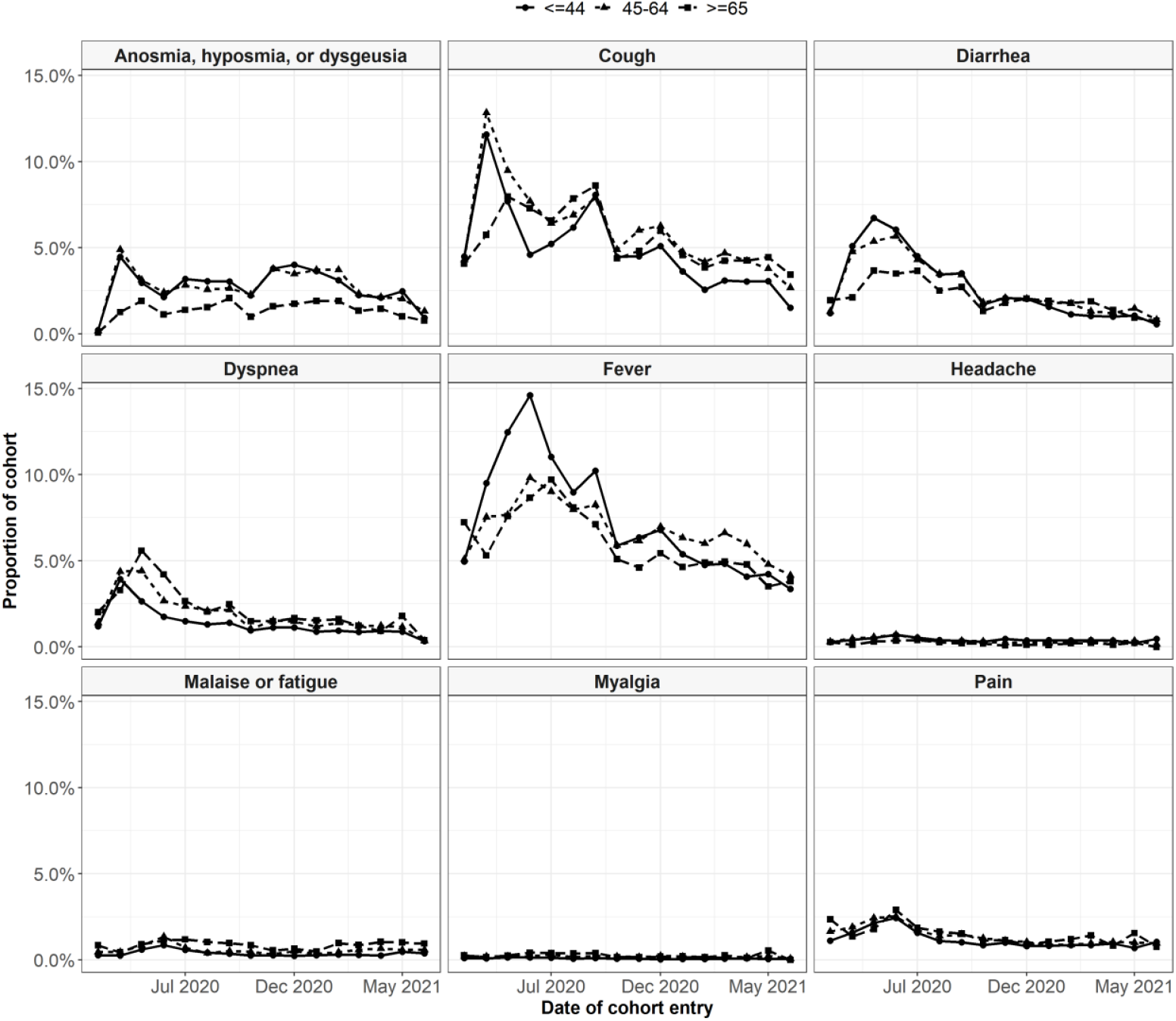
Symptoms recorded at time of outpatient COVID-19 diagnosis or positive test, stratified by age group

**Supplementary Figure 4.**
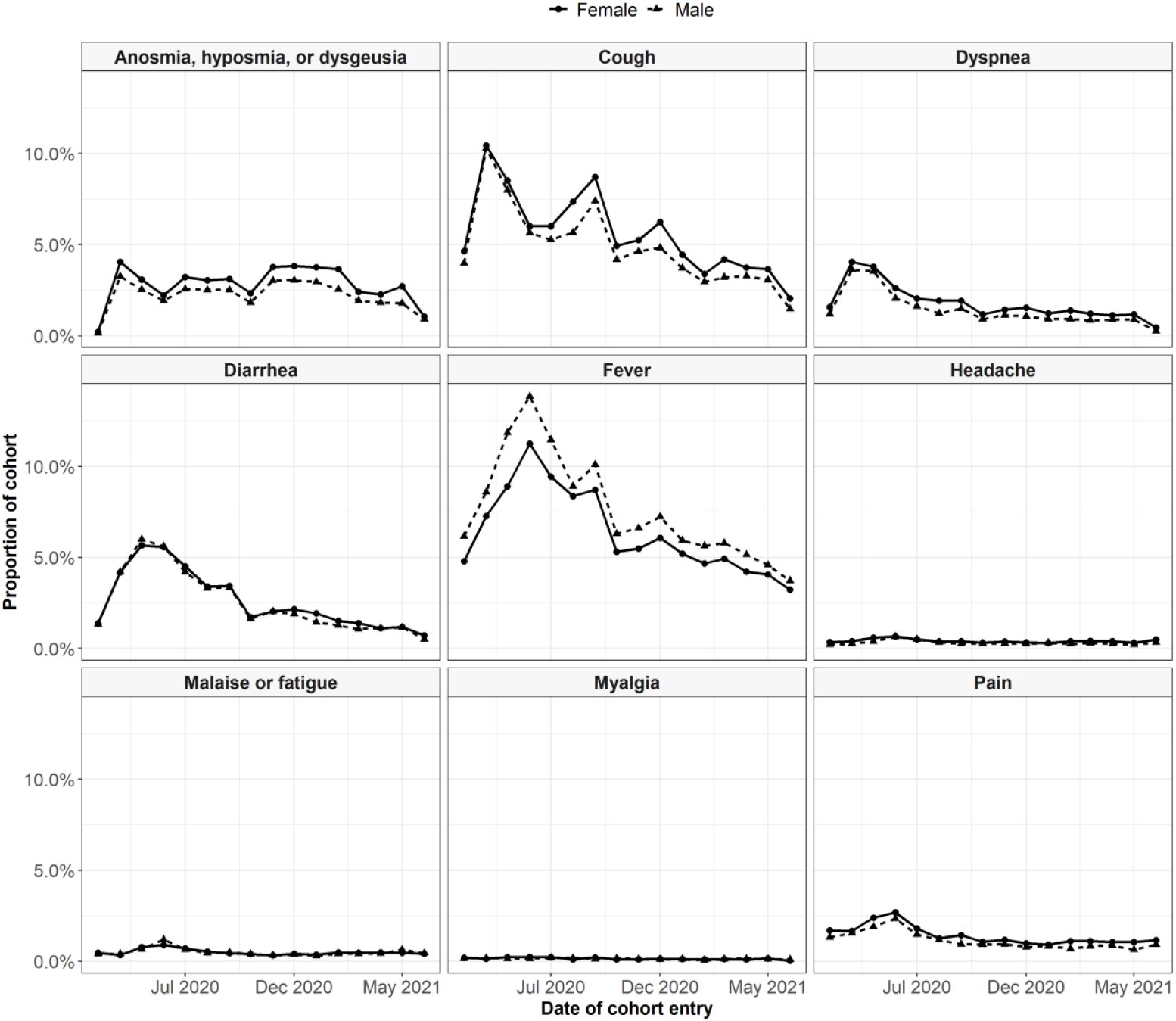
Symptoms recorded at time of outpatient COVID-19 diagnosis or positive test, stratified by sex

